# First-24-hour machine learning for 30-day mortality prediction in ICU trauma patients: development in MIMIC-III and cross-database evaluation in MIMIC-IV

**DOI:** 10.64898/2026.07.28.26359155

**Authors:** Nausin Kudrot, Yong Si, Janet Sanjaya, Sakshie Pathak, Mohammadsaeed Haghi, Kamiar Alaei, Greg Placencia, Maryam Pishgar

## Abstract

ICU trauma patients are clinically heterogeneous, and early mortality risk stratification may support monitoring and resource allocation. We developed machine learning models for 30-day mortality prediction using information recorded during the first 24 hours after ICU admission. In MIMIC-III, six feature configurations were trained using 3,411 patients and compared in a patient-level configuration-selection hold-out subset of 853 patients. The selected XGBoost configuration yielded an area under the precision–recall curve (AUPRC) of 0.556 and an area under the receiver operating characteristic curve (AUROC) of 0.863. For cross-database evaluation, a 228-predictor harmonized XGBoost model was refitted on the complete MIMIC-III cohort and evaluated in 13,747 MIMIC-IV ICU stays without using MIMIC-IV outcomes for model development or recalibration. It achieved an AUPRC of 0.495, an AUROC of 0.825, and a Brier score of 0.109. Calibration was monotonic but showed increasing overprediction at higher predicted risks. First-24-hour clinical information retained predictive value across MIMIC database versions, although internal configuration selection, model differences, same-center provenance, and incomplete feature-mapping documentation limit generalizability and deployment readiness.

## 1 Introduction

Trauma remains a major cause of mortality and disability worldwide and is a frequent reason for admission to the intensive care unit (ICU). Injuries were responsible for an estimated 4.3 million deaths globally in 2019, while road traffic injuries alone cause approximately 1.19 million deaths annually and remain the leading cause of death among individuals aged 5–29 years [1, 2]. The burden of trauma extends beyond acute mortality to long-term disability, prolonged hospitalization, and substantial health-care utilization. As advances in injury prevention, emergency care, and resuscitation have improved survival after severe injury, increasing attention has shifted toward the early recognition and management of physiologic deterioration among trauma patients admitted to the ICU.

Risk stratification in this population remains challenging because ICU trauma patients are clinically heterogeneous. Some patients deteriorate because of hemorrhagic shock, whereas others experience traumatic brain injury, respiratory failure, sepsis, renal hypoperfusion, or multiorgan dysfunction [3, 4]. Early identification of patients at high risk of death may support closer monitoring, timely escalation of care, and more effective allocation of critical care resources [5, 6]. However, mortality is determined by complex and evolving interactions among neurologic status, hemodynamic instability, oxygenation and ventilatory burden, laboratory abnormalities, and responses to treatment during the first hours of ICU care [3, 7]. Conventional severity scores capture important components of this burden, but may not fully represent the nonlinear interactions and short-term physiologic trajectories that characterize early deterioration after trauma [3, 8].

The increasing availability of electronic health record databases has created opportunities to develop machine learning models for mortality prediction in critically ill trauma populations. MIMIC-III and MIMIC-IV provide detailed patient-level information, including demographics, vital signs, laboratory measurements, interventions, and clinical outcomes [9, 10]. Machine learning methods may complement conventional prediction approaches by identifying nonlinear relationships and interactions within these routinely collected data. Yang et al. developed models for predicting 90-day mortality among ICU trauma patients in MIMIC-III and reported that XGBoost achieved the strongest performance among nine classifiers, with SHAP used to identify influential predictors [11]. Recent studies have also applied interpretable machine-learning approaches to mortality prediction in geriatric traumatic brain injury and post-cardiac-arrest ICU cohorts using MIMIC data [12, 13]. Nevertheless, most ICU machine learning studies remain limited to internal validation, and performance commonly declines when models are evaluated under changes in patient populations, clinical practice, or data structure [14].

Several methodological limitations therefore remain in the existing trauma mortality literature. Some models target longer-term outcomes or incorporate variables that are unavailable at the intended prediction time, including hospital or ICU length of stay. Such variables encode information from the subsequent clinical course and may introduce temporal leakage, thereby overstating the usefulness of a model for early decision support [15]. In addition, early ICU data are often represented using isolated or static measurements, despite the potential prognostic importance of physiologic extremes, variability, and short-term trends. Evaluation frequently emphasizes accuracy or the area under the receiver operating characteristic curve (AUROC), whereas the area under the precision–recall curve (AUPRC) may provide a more informative assessment when mortality is relatively uncommon [16]. Calibration and interpretability also require explicit evaluation because strong discrimination alone does not establish the reliability of predicted probabilities or clarify the clinical signals underlying model predictions [17, 18]. Finally, cross-database validation remains uncommon, limiting evidence that model performance is transportable beyond the original development dataset [14].

Therefore, we developed a first-24-hour machine learning framework for predicting 30-day mortality among ICU trauma patients. MIMIC-III was used for model development and internal feature-configuration selection, while a harmonized MIMIC-IV cohort was used for cross-database evaluation. All predictors were restricted to information recorded during the first 24 hours after ICU admission, and post-window variables such as hospital and ICU length of stay were excluded to reduce temporal leakage. Clinically motivated feature groups incorporated neurologic, respiratory, hemodynamic, renal, metabolic, and hematologic information together with temporal summaries of early physiologic trajectories. Six candidate feature configurations were compared using AUPRC as the primary metric. For cross-database evaluation, a model based on features available in both MIMIC-III and MIMIC-IV was refitted using the complete MIMIC-III cohort and subsequently evaluated in MIMIC-IV. Model performance was assessed using discrimination, calibration, and SHAP-based interpretation. This design examined whether predictive signals derived from first-24-hour clinical information remained informative across changes in clinical era, database schema, documentation practices, and feature availability.

## 2 Methods

### 2.1 Study design and data sources

This retrospective cohort study used the Medical Information Mart for Intensive Care III (MIMIC-III) database for model development and internal evaluation, and the Medical Information Mart for Intensive Care IV (MIMIC-IV) database for cross-database transportability evaluation [9, 10]. Both databases contain de-identified electronic health record data from patients treated at Beth Israel Deaconess Medical Center, but represent different clinical periods and database structures. Accordingly, evaluation in MIMIC-IV was intended to assess transportability across database versions, clinical eras, documentation practices, and feature definitions rather than validation in a fully independent hospital system.

The analysis comprised two related modeling stages. First, six candidate feature configurations were developed using an 80:20 patient-level split of the MIMIC-III cohort. The training subset was used for model fitting and threshold selection, while the hold-out subset was used to compare the candidate feature configurations. Second, predictors available in both database versions were harmonized, and a separate cross-database model was refitted using the complete MIMIC-III cohort before evaluation in MIMIC-IV. MIMIC-IV outcomes were not used for feature selection, hyperparameter selection, model fitting, threshold selection, or recalibration.

Both databases are available through PhysioNet [19]. Access was obtained after completion of the required research training and data use agreement. MIMIC-III and MIMIC-IV contain de-identified data collected under institutional review board approval, with informed-consent requirements waived for secondary analysis of the deidentified records. The study investigators had no access to direct patient identifiers. All analyses were conducted in accordance with the relevant institutional guidelines and regulations.

### 2.2 Cohort definition and outcome

Adult ICU patients aged 18–89 years with trauma-related diagnosis codes associated with the hospitalization were eligible for inclusion. Trauma was identified from hospitalization-level diagnosis records rather than diagnoses explicitly timestamped during the first 24 hours. In the documented MIMIC-IV extraction, the first ICU stay within each hospital admission was retained. The saved MIMIC-III analytic dataset contained one record per patient. Patients were excluded if age or sex information was unavailable or if the ICU stay lasted less than 24 hours. The trauma diagnosis-code criteria are summarized in Supplementary Table S2.

Because predictor variables were summarized over the first 24 hours after ICU admission, the intended prediction time was the end of this 24-hour observation window. The resulting target population therefore comprised patients who remained in the ICU for at least 24 hours. The binary outcome was death within 30 days of ICU admission. Patients who died within 30 days were assigned to the positive class, and all remaining patients were assigned to the negative class.

The MIMIC-III cohort was used for model training and internal feature-configuration selection. The MIMIC-IV cohort was constructed using the available trauma cohort logic and was subsequently harmonized to the MIMIC-III feature schema for cross-database evaluation. The internal MIMIC-III split was performed at the patient level to prevent the same patient from appearing in both the training and configuration-selection subsets. Cohort sizes and 30-day mortality frequencies are summarized in Supplementary Table S1.

### 2.3 Prediction framework and leakage control

The study was formulated as a supervised binary classification task. For each eligible ICU stay, the predictor vector was constructed exclusively from information recorded during the first 24 hours after ICU admission. The model generated a probability of death within 30 days of ICU admission, with binary classifications obtained by applying a decision threshold selected during model development.

The first 24 hours served as a fixed observation window, and the intended prediction time was therefore the end of this window. Variables that required knowledge of the subsequent clinical course were excluded, including hospital length of stay, ICU length of stay, discharge disposition, death-time-derived variables, and other post-window information. This restriction was imposed to reduce temporal leakage and to ensure that the model relied only on information available at the intended prediction time [15].

For the internal MIMIC-III analysis, the cohort was divided at the patient level using an 80:20 split and a random seed of 42. Data-dependent preprocessing operations were fitted using the MIMIC-III training subset and applied to the configuration-selection hold-out subset. For the cross-database analysis, preprocessing was refitted using the complete MIMIC-III cohort restricted to the harmonized feature space and was subsequently applied to MIMIC-IV without re-estimating preprocessing parameters from MIMIC-IV. The operating threshold used in the internal analysis was selected from out-of-fold predictions generated within the MIMIC-III training subset.

Missingness and measurement frequency were interpreted cautiously because missing clinical measurements in electronic health records are not necessarily random [20]. A laboratory test may be absent because a patient is clinically stable, whereas frequent measurement may indicate greater physiologic instability or clinical concern. Measurement-count features were therefore treated as potential markers of both patient acuity and clinical workflow rather than as direct physiologic measurements.

### 2.4 Feature construction

Predictors were organized into clinically interpretable feature families comprising demographics, neurologic status, respiratory status, hemodynamics, renal function and urine output, metabolic and acid–base markers, hematologic and coagulation variables, treatment-intensity indicators, temporal summaries, and selected physiologically motivated ratios or interactions. This organization was used to evaluate whether progressively expanded representations of early clinical status provided additional predictive information.

For repeatedly measured variables, values recorded during the first 24 hours were summarized using complementary temporal statistics. Mean values represented the overall physiologic level; minimum and maximum values captured extreme abnormalities; standard deviation represented within-window variability; first and last values described the beginning and end of the observation period; and change and slope features represented short-term evolution. Measurement counts were included as indicators of monitoring intensity.

Let 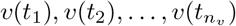 denote the *n*_*v*_ observations of variable *v* recorded at times 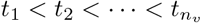 during the first 24 hours. Temporal features were calculated as

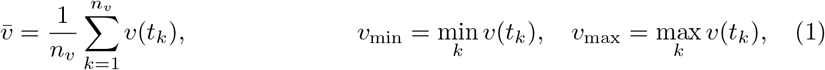

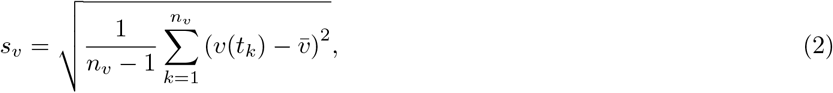

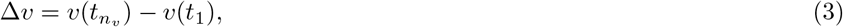

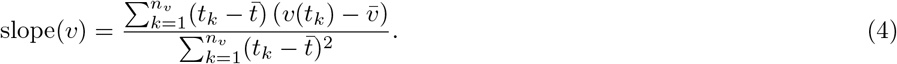

Here, *s*_*v*_ represents within-window variability, Δ*v* represents the net change between the first and last recorded measurements, and slope(*v*) represents the linear trend in the variable over the observation window. For variables with insufficient repeated measurements, temporal statistics requiring multiple observations were treated as missing and handled by the fitted preprocessing pipeline.

Selected ratios and interaction terms were included when they represented clinically plausible relationships between physiologic systems. Examples included

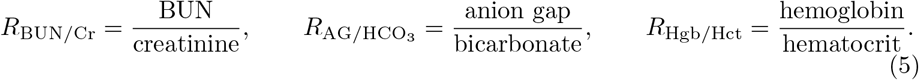

Treatment-intensity indicators, including mechanical ventilation and vasopressor exposure, were also considered together with corresponding respiratory or hemodynamic measurements. These features were intended to distinguish isolated abnormalities from abnormalities occurring despite organ support or active treatment. All features were anchored to ICU admission and restricted to the first 24-hour observation window. Candidate feature configurations ranged from stricter physiologic representations to expanded temporal representations containing additional summaries, ratios, treatment indicators, and contextual variables. Their comparative performance was assessed through the feature-configuration comparison described below.

### 2.5 Internal model development and feature-configuration selection

XGBoost was used as the primary prediction algorithm because the data consisted of heterogeneous, nonlinear, and partially missing tabular electronic health record variables [21]. Gradient-boosted decision trees can capture nonlinear thresholds and interactions among clinical variables without requiring linear relationships or normally distributed predictors.

The MIMIC-III cohort was divided using a patient-level 80:20 split with a random seed of 42, yielding 3,411 patients in the training subset and 853 patients in the configuration-selection hold-out subset. The mortality rates were 13.66% and 13.48%, respectively.

Continuous variables were imputed using the median estimated from the fitting subset. Categorical variables were imputed using the most frequent category and encoded using one-hot encoding, with previously unseen categories ignored during transformation. No feature scaling or normalization was applied. Temporal slope, standard-deviation, and change variables had been precomputed in the supplied analytic feature tables; when these values were unavailable, they were handled as missing values by the fitted preprocessing pipeline.

Six candidate feature configurations were fitted using the MIMIC-III training subset and compared directly in the 853-patient hold-out subset. The all-expanded temporal configuration was selected because it achieved the highest AUPRC in this comparison. Because the same hold-out subset was used both to compare feature configurations and to report the performance of the selected configuration, the resulting estimates represent configuration-selection performance rather than performance in a fully independent test set.

The XGBoost specification used 1,100 estimators, a maximum tree depth of 5, a learning rate of 0.018, row subsampling of 0.78, column subsampling of 0.72, a minimum child weight of 8, L1 regularization of 0.5, L2 regularization of 8.0, and a minimum split-loss reduction of 0.5. The histogram tree-building method, binary logistic objective, and log-loss evaluation metric were used. The random seed was 42. No class weighting, scale-positive weighting, or early stopping was used in the internal feature-configuration analysis. Complete model settings for the internal and harmonized analyses are provided in Supplementary Table S3.

The operating threshold of 0.3282 was selected by maximizing the F1 score using out-of-fold predictions generated within the MIMIC-III training subset. The configuration-selection hold-out subset was not used for threshold selection.

### 2.6 Performance measures

The area under the precision–recall curve (AUPRC) was designated as the primary performance measure because the mortality outcome was imbalanced. Unlike metrics dominated by the majority class, the precision–recall curve directly characterizes the trade-off between the proportion of predicted deaths that are correct and the proportion of observed deaths that are identified [16, 22]. The mortality prevalence in each evaluation cohort was also reported as the no-skill reference value for AUPRC.

The area under the receiver operating characteristic curve (AUROC) was reported as a secondary threshold-independent measure of discrimination and to facilitate comparison with previous mortality-prediction studies. For the internal MIMIC-III analysis, performance at the fixed operating threshold was further characterized using sensitivity, specificity, positive predictive value (PPV), negative predictive value (NPV), F1 score, and the complete confusion matrix. Threshold-dependent MIMIC-IV results were treated as exploratory.

Sensitivity, specificity, PPV, NPV, and F1 score were defined as

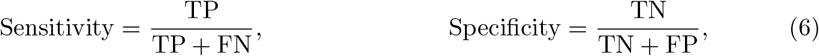

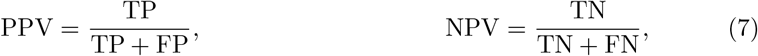

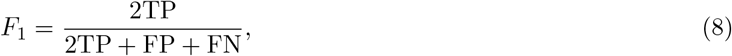

where TP, TN, FP, and FN denote true-positive, true-negative, false-positive, and false-negative classifications, respectively.

Probability calibration was evaluated using calibration plots and the Brier score [17, 23, 24]. The Brier score was calculated as

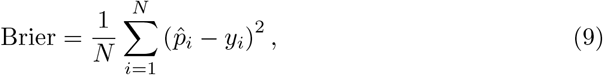

where 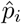 is the predicted probability of 30-day mortality, *y*_*i*_ is the observed binary outcome, and *N* is the number of evaluated analytic records. Lower Brier scores indicate smaller overall probability-prediction error, although the score is influenced by outcome prevalence.

Calibration curves were constructed using 10 equal-frequency bins. Confidence intervals for AUPRC and AUROC were estimated using 1,000 percentile-bootstrap resamples of analytic records. For MIMIC-III, each analytic record represented one patient. For MIMIC-IV, resampling was performed at the ICU-stay level because the documented cohort-construction workflow retained the first ICU stay within each hospital admission. These intervals quantify sampling variability within the evaluated cohort but do not account for optimism introduced by selecting the feature configuration using the same MIMIC-III hold-out subset.

### 2.7 Cross-database transportability evaluation

The internal MIMIC-III feature space contained 259 predictors. For cross-database evaluation, this feature space was restricted to 228 predictors that could be represented in both MIMIC-III and MIMIC-IV. A new XGBoost model was then fitted on the complete MIMIC-III cohort using these harmonized predictors.

The prespecified XGBoost architecture and hyperparameters were retained, while class imbalance was addressed using a scale-positive-weight value equal to the ratio of negative to positive cases in the complete MIMIC-III cohort. Preprocessing parameters were estimated from the complete MIMIC-III fitting cohort and applied to MIMIC-IV without re-estimation. MIMIC-IV outcomes were not used for feature selection, hyperparameter selection, model fitting, or recalibration. A stepwise summary of internal model development and cross-database evaluation is provided in Supplementary Algorithm S1.

Cross-database performance was evaluated using AUPRC, AUROC, the Brier score, and calibration plots. Calibration curves were constructed using 10 equal-frequency bins. Because the internal and harmonized models differed in feature space, training sample, and class weighting, comparisons between their performance estimates were treated as descriptive and were not attributed exclusively to dataset shift.

The operating threshold of 0.3282 had been selected for the unweighted 259-feature internal model. Because the harmonized model used a reduced feature space and class weighting, this threshold was not considered a validated clinical operating point for the MIMIC-IV analysis. Threshold-dependent MIMIC-IV results were therefore treated as exploratory and were not used as primary evidence of transportability.

### 2.8 Model interpretation

SHapley Additive exPlanations (SHAP) were used to examine the contribution of individual predictors to the final XGBoost model [18]. SHAP values quantify the change in model output attributable to each feature relative to the model’s expected prediction. For tree-based models, these values were computed using the TreeSHAP procedure [25].

Global feature importance was summarized using the mean absolute SHAP value across patients in the MIMIC-III configuration-selection hold-out subset:

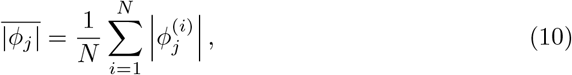

where 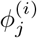 denotes the SHAP value of feature *j* for patient *i*. A global importance plot was used to rank features according to their average contribution, while a SHAP summary plot was used to display both the magnitude and direction of feature contributions.

SHAP analysis was interpreted as an explanation of the associations learned by the model rather than evidence of causal relationships. Particular attention was given to whether highly ranked variables represented clinically plausible early trauma physiology or possible proxies for measurement intensity, coding, and documentation workflow.

### 2.9 Use of generative artificial intelligence

During manuscript preparation, ChatGPT (OpenAI) was used to assist with editorial restructuring and refinement of the manuscript text, and Claude (Anthropic) was used for language refinement. All scientific interpretations, methodological descriptions, and numerical results were independently reviewed and verified by the authors.

## 3 Results

### 3.1 Study cohorts

The MIMIC-III analytic cohort included 4,264 patients, with an overall 30-day mortality rate of 13.63%. The patient-level 80:20 split yielded 3,411 patients in the training subset and 853 patients in the configuration-selection hold-out subset. The corresponding mortality rates were 13.66% and 13.48%, respectively. The hold-out subset included 115 deaths within 30 days of ICU admission.

The harmonized MIMIC-IV cohort included 13,747 ICU stays and 2,175 deaths within 30 days, corresponding to an event rate of 15.82%. For cross-database evaluation, 228 predictors that could be represented in both MIMIC-III and MIMIC-IV were retained, compared with 259 predictors in the internal MIMIC-III feature space.

### 3.2 Internal performance of the selected feature configuration

Six candidate feature configurations were trained using the MIMIC-III training subset and compared directly in the 853-patient configuration-selection hold-out subset. The all-expanded temporal configuration achieved the highest AUPRC and was selected for subsequent internal characterization. Complete point estimates and bootstrap confidence intervals for all six feature configurations are provided in Supplementary Table S4.

In the configuration-selection hold-out subset, the selected model achieved an AUPRC of 0.5563 (95% CI, 0.4720–0.6436) and an AUROC of 0.8633 (95% CI, 0.8306–0.8941). The AUPRC substantially exceeded the mortality-prevalence reference value of 0.1348. The Brier score was 0.0857. The complete performance results are summarized in Table 2.

**Table 1.** Feature families and their clinical rationale.

| Feature family | Examples | Clinical rationale |
| --- | --- | --- |
| Demographics | Age, sex, admission and care context | Represents physiologic reserve and baseline vulnerability following injury. |
| Neurologic status | Glasgow Coma Scale mean, maximum, last value, and slope | Captures the severity and early trajectory of neurologic impairment. |
| Respiratory status | Respiratory-rate summaries, oxygen saturation, mechanical ventilation | Represents respiratory stress, impaired oxygenation, airway protection, and ventilatory support. |
| Hemodynamics | Mean arterial pressure, systolic and diastolic blood pressure, heart-rate trends, shock-index-related features | Captures hypotension, compensatory tachycardia, bleeding, and shock physiology. |
| Renal function and urine output | Blood urea nitrogen, creatinine-related features, 24-hour urine output | Reflects renal perfusion, organ dysfunction, and response to resuscitation. |
| Metabolic and acid-base status | Lactate, bicarbonate, anion gap, glucose, sodium | Represents tissue hypoperfusion, metabolic stress, and acid-base derangement. |
| Hematologic and coagulation status | Hemoglobin, hematocrit, red-cell distribution width, platelets, INR, PT, PTT | Reflects bleeding burden, oxygen-carrying capacity, inflammation, and coagulopathy. |
| Temporal summaries | Mean, minimum, maximum, standard deviation, first, last, change, slope, and measurement count | Captures severity, instability, persistence, and short-term physiologic trajectory. |
| Ratios and interactions | Blood urea nitrogen-to-creatinine ratio, anion-gap-to-bicarbonate ratio, hemoglobin-to-hematocrit ratio, treatment-physiology interactions | Represents coupled pathophysiologic states that may not be captured by isolated variables. |

**Table 2.** Performance of the selected all-expanded temporal configuration in the MIMIC-III configuration-selection hold-out subset.

| Measure | Value |
| --- | --- |
| Number of patients | 853 |
| Deaths within 30 days | 115 |
| Mortality prevalence | 0.1348 |
| AUPRC (95% CI) | 0.5563 (0.4720–0.6436) |
| AUROC (95% CI) | 0.8633 (0.8306–0.8941) |
| Brier score | 0.0857 |
| F1 score | 0.5064 |
| Sensitivity | 0.5130 |
| Specificity | 0.9201 |
| Positive predictive value | 0.5000 |
| Negative predictive value | 0.9238 |
| Confusion matrix | TN = 679, FP = 59, FN = 56, TP = 59 |

At the operating threshold of 0.3282, which had been selected by maximizing the F1 score using out-of-fold predictions generated within the MIMIC-III training subset, the model correctly classified 59 of the 115 deaths and 679 of the 738 survivors. This corresponded to a sensitivity of 51.3%, specificity of 92.0%, positive predictive value of 50.0%, negative predictive value of 92.4%, and F1 score of 0.5064.

The calibration plot showed generally close agreement between predicted and observed mortality across the lower and intermediate predicted-risk ranges, while the highest displayed risk group showed some overprediction (Figure 1). The Brier score of 0.0857 indicated relatively low overall probability-prediction error within this subset.

**Fig. 1.**
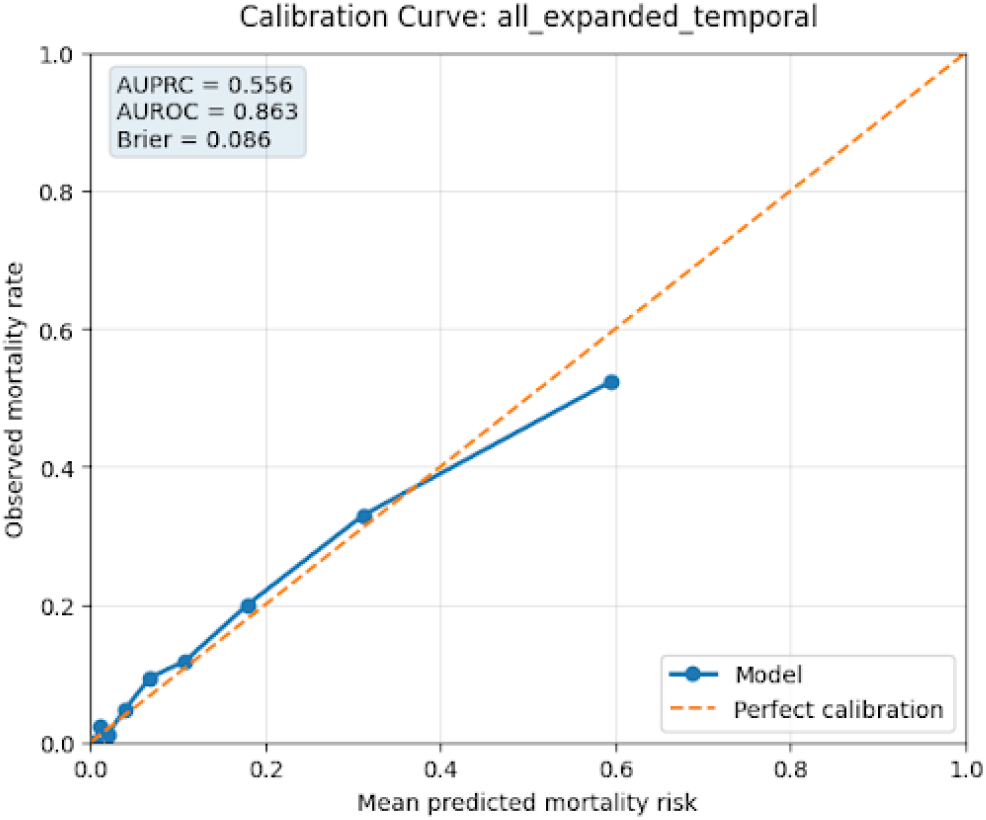
Calibration of predicted 30-day mortality probabilities for the selected configuration in the MIMIC-III configuration-selection hold-out subset. The diagonal line represents perfect agreement between predicted and observed mortality.

**Fig. 2.**
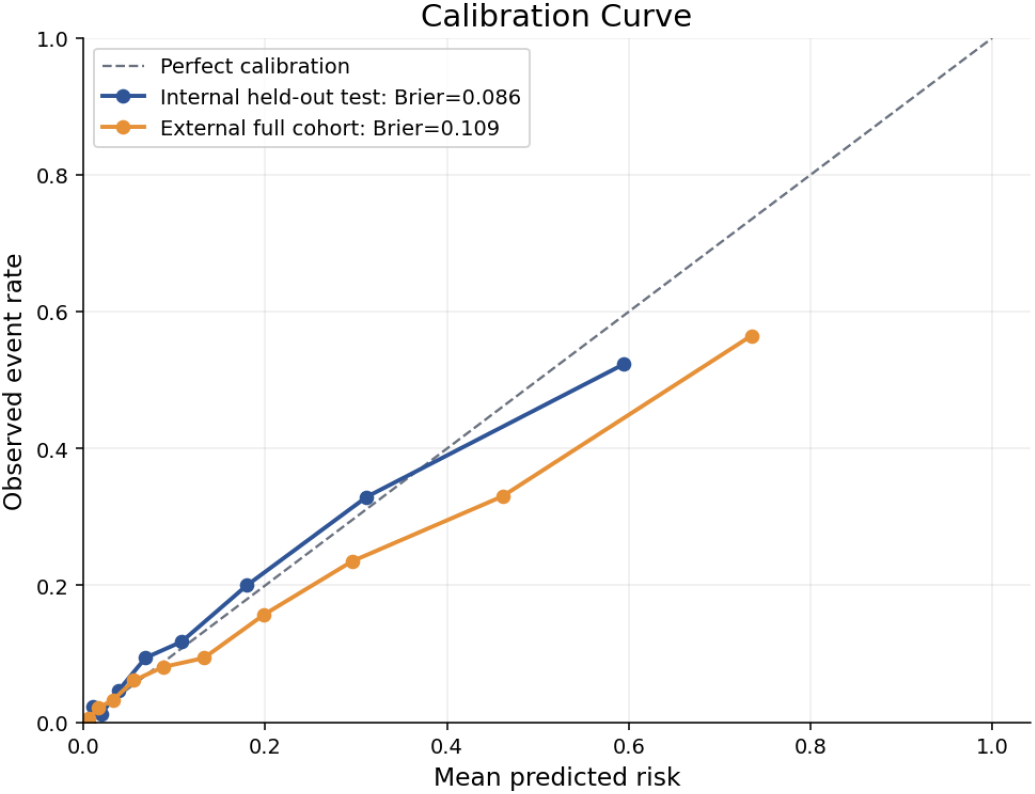
Calibration of predicted 30-day mortality probabilities in the MIMIC-III configuration-selection hold-out subset and the MIMIC-IV cross-database evaluation cohort. Calibration curves were constructed using 10 equal-frequency bins. The diagonal line represents perfect agreement between predicted and observed mortality.

Because the same hold-out subset was used both to compare the six feature configurations and to report the performance of the selected configuration, these results represent configuration-selection performance rather than performance in a fully independent test set. The reported bootstrap confidence intervals quantify sampling variability within this subset but do not account for potential optimism introduced by feature-configuration selection.

### 3.3 Feature-configuration analysis

Performance was broadly consistent across the six feature configurations evaluated in the MIMIC-III configuration-selection hold-out subset. AUPRC ranged from 0.5235 to 0.5563, while AUROC ranged from 0.8633 to 0.8691 (Table 3). The all-expanded temporal configuration achieved the highest AUPRC at 0.5563 and was selected on this basis, whereas the all-expanded temporal v2 configuration achieved the highest AUROC at 0.8691.

**Table 3.** Performance of the evaluated feature configurations in the MIMIC-III configuration-selection hold-out subset.

| Feature configuration | AUPRC | AUROC |
| --- | --- | --- |
| strict_temporal_full | 0.5235 | 0.8655 |
| strict_temporal_full_v2 | 0.5249 | 0.8642 |
| aggressive_best_rebuild | 0.5424 | 0.8663 |
| aggressive_best_rebuild_v2 | 0.5276 | 0.8653 |
| all_expanded_temporal_v2 | 0.5516 | 0.8691 |
| all_expanded_temporal | **0.5563** | 0.8633 |

The relatively narrow range of AUROC values indicates that all configurations retained similar ranking performance. Expanded temporal representations produced modestly higher AUPRC values than the strict temporal configurations. However, these comparisons were performed in the same subset used to select the reported configuration, and the observed differences should therefore be interpreted descriptively rather than as independently validated improvements.

### 3.4 Cross-database evaluation in MIMIC-IV

For cross-database evaluation, the 259-predictor internal MIMIC-III feature space was restricted to 228 predictors that could be represented in both MIMIC-III and MIMIC-IV. A separate harmonized XGBoost model was then refitted using the complete MIMIC-III cohort of 4,264 patients. The prespecified model architecture and hyperparameters were retained, while class imbalance was addressed by setting `scale _pos_weight` to the ratio of negative to positive cases in the complete MIMIC-III cohort.

The preprocessing pipeline was refitted using the complete MIMIC-III cohort within the harmonized feature space and subsequently applied to MIMIC-IV without re-estimating preprocessing parameters from MIMIC-IV. MIMIC-IV outcomes were not used for feature selection, hyperparameter selection, model fitting, or recalibration.

The harmonized model was evaluated in 13,747 MIMIC-IV ICU stays, including 2,175 deaths within 30 days of ICU admission, corresponding to a mortality prevalence of 15.82%. The model achieved an AUPRC of 0.4952 (95% CI, 0.473–0.519), compared with the prevalence reference value of 0.1582, and an AUROC of 0.8249 (95% CI, 0.816–0.834). The Brier score was 0.109.

A descriptive comparison of the internal configuration-selection results and the MIMIC-IV cross-database results is presented in Table 4.

**Table 4.** Descriptive comparison of internal configuration-selection performance and cross-database performance in MIMIC-IV.

| Analysis | AUPRC (95% CI) |  | AUROC (95% CI) |  | Brier score |
| --- | --- | --- | --- | --- | --- |
| Selected internal configuration in the MIMIC-III configuration-selection hold-out subset | 0.5563 | (0.4720–0.6436) | 0.8633 | (0.8306–0.8941) | 0.0857 |
| Harmonized model evaluated in the MIMIC-IV cross-database cohort | 0.4952 | (0.473–0.519) | 0.8249 | (0.816–0.834) | 0.109 |

Calibration in MIMIC-IV remained approximately monotonic across increasing predicted-risk groups, but the model increasingly overestimated mortality at higher predicted probabilities. In the highest predicted-risk decile, the mean predicted probability was 0.7352, whereas the observed 30-day mortality rate was 0.5651. Smaller differences between predicted and observed risk were present in the lower-risk groups, while overprediction became more apparent from the sixth risk decile onward. The complete calibration-bin results are provided in Supplementary Table S5.

The calibration curves were constructed using 10 equal-frequency bins. Because the harmonized model incorporated class weighting and was not recalibrated after fitting, its probability outputs should not be interpreted as directly calibrated estimates of absolute mortality risk in MIMIC-IV.

The internal and cross-database performance estimates should be compared cautiously because they were generated using related but non-identical models. The internal analysis used 259 predictors, was trained on 3,411 patients without class weighting, and was selected based on performance in the 853-patient configuration-selection subset. In contrast, the cross-database analysis used 228 harmonized predictors, was refitted using all 4,264 MIMIC-III patients, and incorporated scale-positive weighting. Consequently, the numerical difference between the internal and MIMIC-IV results cannot be attributed exclusively to temporal or database shift.

### 3.5 Model interpretation

SHAP analysis of the selected internal configuration showed that age had the largest mean absolute contribution to predictions in the MIMIC-III configuration-selection hold-out subset, followed by the number of respiratory-rate measurements, maximum Glasgow Coma Scale (GCS) score, 24-hour urine output, respiratory-rate variability, and the last recorded GCS score (Figure 3). Additional influential features included respiratory-rate mean, GCS mean and slope, heart-rate slope, hematologic measurements, lactate, and mean arterial pressure.

**Fig. 3.**
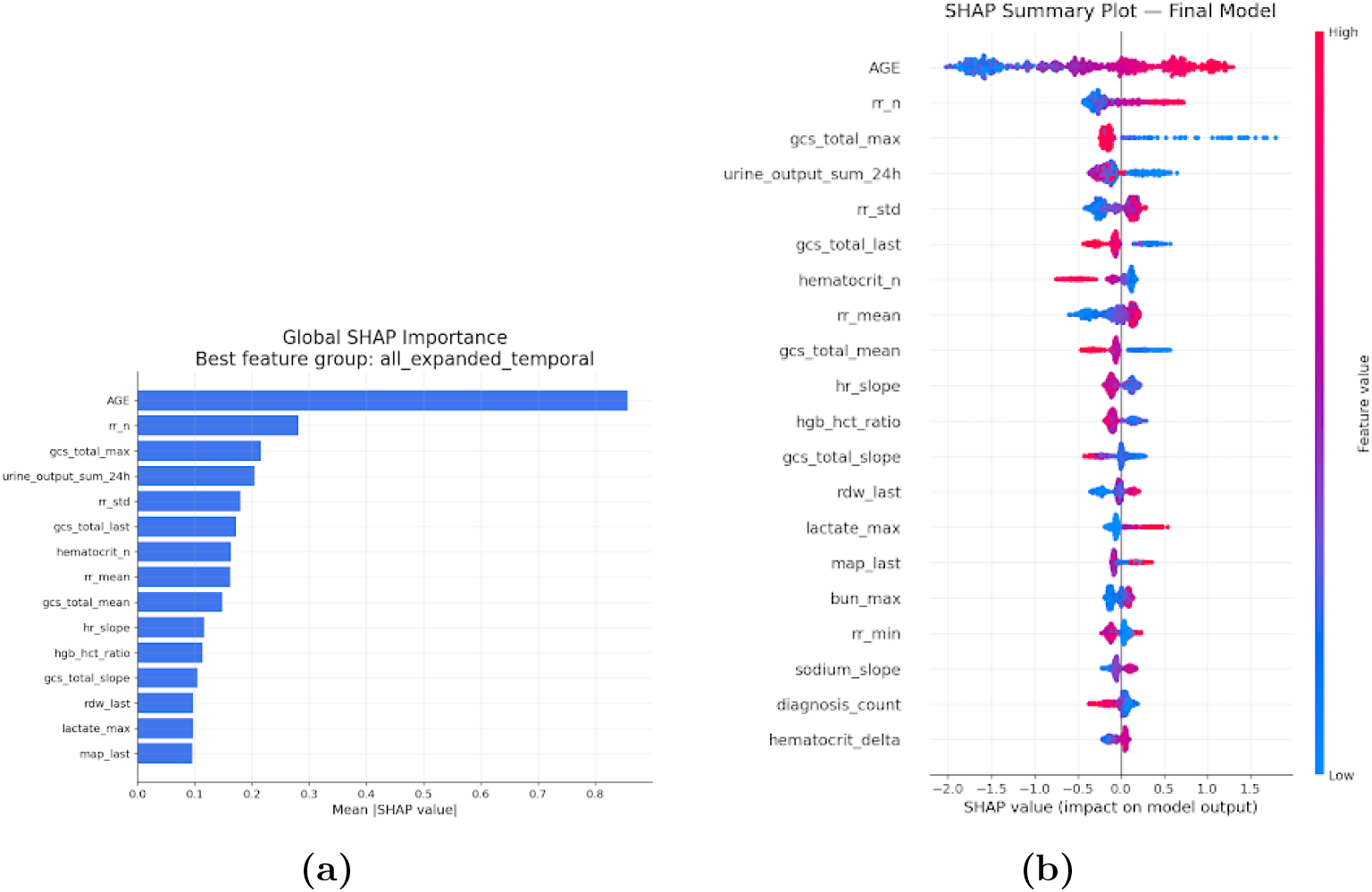
SHAP-based interpretation of the selected XGBoost configuration in the MIMIC-III configuration-selection hold-out subset. **(a)** Global feature importance ranked by mean absolute SHAP value. **(b)** SHAP summary plot showing the magnitude and direction of individual feature contributions. Positive SHAP values increase the predicted probability of 30-day mortality, while negative values decrease it. Feature color represents the relative observed value from low to high.

The SHAP summary plot indicated that older age was consistently associated with higher predicted mortality risk. Lower maximum, mean, and last GCS values were generally associated with positive SHAP values, indicating increased predicted risk among patients with greater neurologic impairment. Lower 24-hour urine output and lower final mean arterial pressure were also associated with higher risk, while elevated lactate and red-cell distribution width contributed positively to mortality predictions.

Respiratory-rate features showed substantial contributions across several representations. Higher respiratory-rate measurement frequency, mean, and variability were generally associated with increased predicted risk. These features may reflect respiratory instability, increased monitoring intensity, or both. Accordingly, measurement-count variables and other documentation-related features were interpreted cautiously because they may capture clinical workflow and clinician concern in addition to underlying physiology.

Overall, the dominant SHAP patterns were compatible with clinically recognizable pathways involving reduced physiologic reserve, neurologic injury, respiratory instability, impaired perfusion, metabolic stress, and hematologic abnormalities. Nevertheless, SHAP values describe associations learned by the model and should not be interpreted as causal effects.

## 4 Discussion

This study evaluated whether clinical information recorded during the first 24 hours after ICU admission could support 30-day mortality prediction among trauma patients. In the internal MIMIC-III analysis, six candidate feature configurations were trained using 3,411 patients and compared in a patient-level configuration-selection hold-out subset of 853 patients. The selected all-expanded temporal configuration achieved an AUPRC of 0.5563 and an AUROC of 0.8633 in this subset. For cross-database evaluation, a separate model based on 228 predictors available in both MIMIC-III and MIMIC-IV was refitted using the complete MIMIC-III cohort and evaluated in 13,747 MIMIC-IV ICU stays. This harmonized model achieved an AUPRC of 0.4952, an AUROC of 0.8249, and a Brier score of 0.109. Taken together, these findings indicate that first-24-hour clinical information contained predictive signal across both MIMIC database versions, while also showing important limitations in internal model selection, feature harmonization, and probability calibration.

The present study extends previous work on machine learning for mortality prediction in ICU trauma populations. Yang et al. reported that XGBoost achieved the strongest performance among nine classifiers for predicting 90-day mortality using MIMIC-III [11]. That study demonstrated the potential of nonlinear tree-based models in this population, but its predictors included hospital and ICU length of stay, which are unavailable at an early prediction landmark and may encode information from the subsequent clinical course. The present analysis instead restricted all predictors to the first 24 hours after ICU admission and excluded length-of-stay, discharge, and other post-window variables to reduce temporal leakage [15]. It also emphasized AUPRC for the imbalanced outcome and assessed a harmonized model in MIMIC-IV. Direct numerical comparison with the earlier 90-day study remains inappropriate because the outcome horizon, cohort construction, feature definitions, model-selection procedures, and evaluation designs differed.

The feature-configuration comparison suggested that expanded temporal information provided a modest improvement in precision–recall performance. AUPRC ranged from 0.5235 to 0.5563 across the six configurations, whereas AUROC varied only from 0.8633 to 0.8691. The all-expanded temporal configuration achieved the highest AUPRC, but the relatively narrow differences indicate that increasing feature complexity did not produce a large improvement in overall discrimination. Temporal summaries may nevertheless contribute clinically relevant information by distinguishing persistent or worsening abnormalities from isolated measurements. For example, patients with similar average blood pressure may have different prognoses depending on whether hypotension resolves or persists, while a declining GCS trajectory may convey information that is not represented by a single neurologic assessment.

These internal comparisons require cautious interpretation because the same 853-patient hold-out subset was used both to compare the six candidate feature configurations and to report the performance of the selected configuration. The resulting estimates therefore represent configuration-selection performance rather than performance in a fully untouched test set and may be optimistic. The percentile-bootstrap confidence intervals quantify sampling variability within this subset, but they do not account for the additional uncertainty introduced by selecting the feature configuration on the basis of performance in the same data. Future development should separate feature selection, hyperparameter selection, and final performance evaluation through nested cross-validation or an additional independent test subset.

The SHAP analysis identified clinically recognizable predictors of mortality risk. Age had the largest overall contribution, consistent with reduced physiologic reserve and increased vulnerability among older trauma patients. GCS summaries represented the severity and short-term trajectory of neurologic impairment, while urine output and mean arterial pressure reflected perfusion and organ function. Lactate, anion gap, hematologic measurements, and heart-rate trends captured aspects of tissue hypoperfusion, metabolic stress, bleeding, and hemodynamic instability. These patterns support the clinical face validity of the selected model, but they should not be interpreted as causal effects. SHAP describes associations used by a fitted prediction model and does not establish that changing an influential variable would alter mortality risk.

Measurement-frequency variables warrant particular caution. The number of respiratory-rate measurements was among the most influential predictors, but measurement frequency may reflect both physiologic instability and process-of-care factors such as clinician concern, monitoring intensity, staffing patterns, and documentation practices. Such variables can improve prediction within a database while being sensitive to local workflows. Their prominence reinforces the importance of cross-database assessment and suggests that future models should distinguish physiologic measurements from workflow-dependent predictors. Future sensitivity analyses using prespecified physiologic-only feature sets would help determine the extent to which predictive performance depends on documentation and care-process variables.

The MIMIC-IV findings provide evidence that harmonized first-24-hour predictors remained informative across differences in clinical period and database structure. However, the internal and cross-database results were generated using related but non-identical model specifications. The internal analysis used 259 predictors, was trained on 3,411 patients without class weighting, and selected its feature configuration using the 853-patient hold-out subset. In contrast, the MIMIC-IV analysis used 228 harmonized predictors, was refitted using all 4,264 MIMIC-III patients, and incorporated scale-positive weighting. Consequently, the decrease from an internal AUPRC of 0.5563 and AUROC of 0.8633 to a MIMIC-IV AUPRC of 0.4952 and AUROC of 0.8249 cannot be attributed exclusively to temporal or database shift. Differences in feature space, training sample size, and class weighting also contributed to the change in performance.

Calibration analysis further limits claims of deployment readiness. In MIMIC-IV, observed mortality increased monotonically across predicted-risk groups, indicating that the model retained value for relative risk ranking. However, predicted probabilities increasingly exceeded observed mortality at higher risk levels. In the highest predicted-risk decile, the mean predicted probability was 0.7352, compared with an observed mortality rate of 0.5651. The external Brier score was 0.109. This overprediction is not unexpected because the harmonized model incorporated class weighting and was not recalibrated, but it means that the raw output probabilities should not be interpreted as directly calibrated estimates of absolute mortality risk. Recalibration in the target population would be necessary before probability-based clinical use [17].

Threshold-dependent MIMIC-IV results were not treated as primary evidence because the threshold of 0.3282 was originally selected using out-of-fold predictions from the unweighted 259-feature internal model. The cross-database model used a reduced feature space, a larger fitting cohort, and scale-positive weighting, all of which may alter the probability distribution and appropriate operating threshold. Applying the internal threshold to the harmonized model may be informative as an exploratory analysis, but it does not establish a validated clinical decision point. Any future threshold should be selected using a prespecified clinical objective and independently evaluated in the intended target population. Exploratory threshold-dependent metrics and confusion-matrix counts are reported in Supplementary Tables S6 and S7.

Several additional limitations should be considered. First, this was a retrospective study using routinely collected electronic health record data, and the models identify predictive associations rather than causal mechanisms. Second, trauma eligibility was determined from hospitalization-level diagnosis codes, which may not fully capture injury mechanism, anatomic severity, operative findings, or the timing of individual injuries. The original MIMIC-III cohort-extraction code was unavailable, which limited complete verification that the MIMIC-III and MIMIC-IV cohort definitions were identical. Third, patients who died, recovered, or left the ICU before 24 hours were excluded because a complete first-day observation window was required. The target population therefore represents patients alive and remaining in the ICU at the 24-hour landmark and should not be interpreted as an admission-time cohort.

Fourth, feature harmonization was constrained by differences in item identifiers, database schema, units, measurement practices, and missingness. Although 228 predictors were represented in both databases, a complete feature-mapping dictionary containing source tables, item identifiers, units, and aggregation rules was not available. This limits reproducibility and makes it difficult to determine whether apparently equivalent variables were constructed identically. In addition, the documented MIMIC-IV workflow retained the first ICU stay within each hospital admission but did not document a further restriction to one hospitalization per individual. Repeated hospitalizations from the same individual may therefore remain in the MIMIC-IV cohort, and the external confidence intervals were not estimated using subject-clustered resampling.

Future work should evaluate a prespecified feature configuration using nested model selection and a fully untouched internal test set. The resulting model should then be evaluated prospectively or retrospectively in independent multicenter datasets without changing the feature definition, model specification, or probability scale. A complete harmonization dictionary should document every variable’s source table, item identifier, unit, observation window, and aggregation rule. Future analyses should also evaluate calibration intercept and slope, compare recalibration approaches, assess clinically relevant subgroups, and use decision-curve analysis to determine whether predicted risk provides net benefit across plausible operating thresholds. Incorporating trauma-specific information, including anatomic injury severity, body-region involvement, transfusion burden, operative procedures, and injury mechanism, may further improve prediction while reducing reliance on workflow-sensitive variables.

## 5 Conclusion

This study evaluated first-24-hour machine learning models for predicting 30-day mortality among ICU trauma patients. In MIMIC-III, the selected all-expanded temporal XGBoost configuration achieved an AUPRC of 0.5563 and an AUROC of 0.8633 in an 853-patient hold-out subset used for feature-configuration selection. For cross-database evaluation, a separate 228-feature harmonized model was refitted using the complete MIMIC-III cohort and evaluated in MIMIC-IV, achieving an AUPRC of 0.4952, an AUROC of 0.8249, and a Brier score of 0.109. These findings indicate that clinical information recorded during the first 24 hours retained predictive value across MIMIC database versions.

The results should nevertheless be interpreted as evidence from a research-stage prediction framework rather than a deployment-ready clinical model. Internal performance may be optimistic because the reported subset was used for feature-configuration selection, while the internal and cross-database results were generated using different feature spaces, training samples, and class-weighting strategies. External overprediction at higher risk levels, incomplete feature-mapping documentation, retrospective design, and the same-center provenance of both databases further limit generalizability. Fully independent model selection and testing, reproducible feature harmonization, target-population recalibration, and multicenter validation are required before clinical implementation.

## Data Availability

The MIMIC-III and MIMIC-IV datasets analysed in this study are available through PhysioNet, subject to completion of the required credentialing, research training, and data use agreements. No new patient data were collected. Derived analytic datasets cannot be publicly redistributed because access remains subject to the PhysioNet data use agreements. The model-fitting, evaluation, calibration, and visualization code available to the authors may be obtained from the corresponding author upon reasonable request. Access to and use of the code remain subject to the data-use restrictions applicable to the MIMIC databases.

## Supplementary information

Supplementary Information accompanies this manuscript and includes additional methodological details, cohort and trauma diagnosis-code criteria, complete model settings, feature-configuration comparisons, calibration points, and exploratory threshold-dependent analyses.

## Acknowledgements

The authors thank the Massachusetts Institute of Technology Laboratory for Computational Physiology and Beth Israel Deaconess Medical Center for developing and maintaining the MIMIC databases and making them available through PhysioNet.

## Declarations

### Ethics approval and consent to participate

This study used de-identified data from the MIMIC-III and MIMIC-IV databases. The creation and use of the MIMIC databases were approved by the institutional review boards of Beth Israel Deaconess Medical Center and the Massachusetts Institute of Technology, with the requirement for individual informed consent waived because the data were de-identified. Access to the databases was obtained after completion of the required research training and execution of the applicable data use agreements. The present study involved secondary analysis of de-identified data and no direct interaction with human participants.

### Consent for publication

Not applicable.

### Data availability

The MIMIC-III and MIMIC-IV datasets analysed in this study are available through PhysioNet, subject to completion of the required credentialing, research training, and data use agreements. No new patient data were collected. Derived analytic datasets cannot be publicly redistributed because access remains subject to the PhysioNet data use agreements.

### Materials availability

Not applicable.

### Code availability

The model-fitting, evaluation, calibration, and visualization code available to the authors may be obtained from the corresponding author upon reasonable request. Access to and use of the code remain subject to the data-use restrictions applicable to the MIMIC databases.

### Author contributions

N.K., Y.S., J.S., S.P., and M.H. contributed equally to this work. N.K., J.S., S.P., and M.H. contributed to data curation, feature preparation, model development, validation, visualization, and preparation of the original manuscript draft. Y.S. conducted the methodological assessment, evaluated the consistency of the reported analyses, restructured the manuscript, interpreted the modeling results, and led manuscript revision, writing, review, and editing. K.A. provided clinical interpretation and critically reviewed the manuscript. G.P. contributed to methodological review and manuscript revision. M.P. conceived and supervised the study, coordinated the project, interpreted the results, and critically revised the manuscript. All authors reviewed and approved the final manuscript.

## References

World Health Organization. Injuries and violence (2024). URL https://www.who.int/news-room/fact-sheets/detail/injuries-and-violence. Fact sheet, 19 June 2024.

GBD 2019 Injuries Collaborators. Global, regional, and national burden of injuries, and burden attributable to injuries risk factors, 1990 to 2019: results from the Global Burden of Disease Study 2019. Public Health 237, 212–231 (2024).

Roepke, R. M. L. et al. Predictive performance for hospital mortality of SAPS 3, SOFA, ISS, and New ISS in critically ill trauma patients: a validation cohort study. Journal of Intensive Care Medicine 39, 44–51 (2024).

Majdán, M. et al. Glasgow coma scale motor score and pupillary reaction to predict six-month mortality in patients with traumatic brain injury. Journal of Neurotrauma 32, 101–108 (2015).

Vang, M., Ostberg, M., Steinmetz, J. & Rasmussen, L. S. Shock index as a predictor for mortality in trauma patients: a systematic review and meta-analysis. European Journal of Trauma and Emergency Surgery 48, 2559–2566 (2022).

Carsetti, A. et al. Shock index as predictor of massive transfusion and mortality in patients with trauma: a systematic review and meta-analysis. Critical Care 27, 85 (2023).

Servià, L., Badia, M., Montserrat, N. & Trujillano, J. Severity scores in trauma patients admitted to ICU. physiological and anatomic models. Medicina Intensiva 43, 26–34 (2019).

Girshausen, R. et al. Polytrauma scoring revisited: prognostic validity and usability in daily clinical practice. European Journal of Trauma and Emergency Surgery 49, 393–401 (2023).

Johnson, A. E. W. et al. MIMIC-III, a freely accessible critical care database. Scientific Data 3, 160035 (2016).

Johnson, A. E. W. et al. MIMIC-IV, a freely accessible electronic health record dataset. Scientific Data 10, 1 (2023).

Yang, S., Cao, L., Zhou, Y. & Hu, C. A retrospective cohort study: predicting 90-day mortality for ICU trauma patients with a machine learning algorithm using XGBoost using MIMIC-III database. Journal of Multidisciplinary Healthcare 16, 2625–2640 (2023).

Si, Y. et al. Machine learning-based prediction of mortality in geriatric traumatic brain injury patients. BioMedInformatics 6, 17 (2026).

Si, Y. et al. Retrospective machine learning approach for forecasting in-hospital death in ICU patients after cardiac arrest. Informatics in Medicine Unlocked 64, 101776 (2026).

Rockenschaub, P. et al. External validation of AI-based scoring systems in the ICU: a systematic review and meta-analysis. BMC Medical Informatics and Decision Making 25, 5 (2025).

Davis, S. E., Matheny, M. E., Balu, S. & Sendak, M. P. A framework for understanding label leakage in machine learning for health care. Journal of the American Medical Informatics Association 31, 274–280 (2024).

Saito, T. & Rehmsmeier, M. The precision-recall plot is more informative than the ROC plot when evaluating binary classifiers on imbalanced datasets. PLOS ONE 10, e0118432 (2015).

Van Calster, B., McLernon, D. J., van Smeden, M., Wynants, L. & Steyerberg, E. W. Calibration: the achilles heel of predictive analytics. BMC Medicine 17, 230 (2019).

Lundberg, S. M. & Lee, S.-I. A unified approach to interpreting model predictions. Advances in Neural Information Processing Systems 30, 4765–4774 (2017).

Goldberger, A. L. et al. PhysioBank, PhysioToolkit, and PhysioNet: components of a new research resource for complex physiologic signals. Circulation 101, e215–e220 (2000).

Groenwold, R. H. H. Informative missingness in electronic health record systems: the curse of knowing. Diagnostic and Prognostic Research 4, 8 (2020).

Chen, T. & Guestrin, C. XGBoost: a scalable tree boosting system. Proceedings of the 22nd ACM SIGKDD International Conference on Knowledge Discovery and Data Mining 785–794 (2016).

Davis, J. & Goadrich, M. The relationship between precision-recall and ROC curves. Proceedings of the 23rd International Conference on Machine Learning 233–240 (2006).

Brier, G. W. Verification of forecasts expressed in terms of probability. Monthly Weather Review 78, 1–3 (1950).

Steyerberg, E. W. et al. Assessing the performance of prediction models: a framework for some traditional and novel measures. Epidemiology 21, 128–138 (2010).

Lundberg, S. M. et al. From local explanations to global understanding with explainable AI for trees. Nature Machine Intelligence 2, 56–67 (2020).

